# Concordance of Truvian’s Benchtop Blood Testing Platform to Central Laboratory Testing

**DOI:** 10.1101/2023.11.27.23299081

**Authors:** Reneé Higgins, Nicholas Haase, Patrick Desmond, Ian Levine, Clara Romero, Brian Fernández, Eumene Lee, Mike Adams, Derek Arndt, Greg Grabarek, Ju Young Kim, Rachel Krupa, Ginger Mina, Ryan Morgan, John Poland, Galen Reed, Robin Richardson, Kelline Rodems, Astrid Schroeder, Maike Zimmermann, Florence Lee, Dena Marrinucci

**Affiliations:** Truvian

## Abstract

**Introduction:** Routine blood tests play an essential role in modern healthcare, but their administration, processing, and reporting under the current centralized testing model is slow, inefficient, and cumbersome for patients and providers especially in outpatient settings.

Truvian’s benchtop blood testing platform, in late-stage development, aims to decentralize and streamline routine blood testing, replacing traditional send-outs to a central laboratory with a compact, easy-to-use benchtop platform at the point-of-action^™^ to ensure timely and actionable results between a patient and healthcare provider. Using only a small amount of blood from a single heparinized sample, the Truvian platform can simultaneously provide results for a full panel of routine blood tests spanning clinical chemistry, hematology, and immunoassays.

Evaluation of the Truvian platform in independent external studies is important to understand its performance in real world settings and to identify opportunities for improvement to complete development. To assess the performance of a comprehensive wellness panel on the Truvian platform, a multi-site study was completed at Truvian’s headquarters in San Diego, California, and at an independent clinical trial site in the Pacific Northwest. The study evaluated the panel’s precision and accuracy against central laboratory analyzers.

**Methods:** Precision and accuracy studies were performed with a panel of 32 routine blood tests including immunoassay, clinical chemistry and hematology assays on the Truvian platform. Precision studies were run across multiple days and instruments to assess repeatability and reproducibility for each test in the panel. A method comparison study included 237 patients and compared the Truvian platform to best-in-class FDA cleared central laboratory analyzers – the Roche Cobas and Sysmex analyzers. Bland-Altman and either Passing-Bablok, or Deming regression analyses were used to determine agreement for each analyte in the panel. Additionally, linearity, sensitivity, and endogenous interfering substance studies were carried out for tests within the panel.

**Results:** Overall, the Truvian platform had a run reliability rate of > 95%. In the precision and detection capability studies, the evaluated tests successfully satisfied all predefined criteria for precision, linearity, and sensitivity. Moreover, the method comparison study revealed concordance with central laboratory analyzers.

**Conclusions:** This multi-site study demonstrated that the Truvian platform, currently in late-stage development, is capable of delivering the clinical performance and reliability needed for decentralized blood testing. The study also provided additional areas of focus to complete development.

## INTRODUCTION

Timely access to actionable data from routine blood tests can support wellness, inform medical care, and save lives^1-3^. In fact, 70% of medical decisions are made based on diagnostic testing^4^. These tests provide insight into a person’s immunological, metabolic, and dietary health. Routine blood testing is important for identifying early-stage conditions. With 60% of adults living with at least one chronic disease^5^, testing is important to preserve the quality of life for those already afflicted.

Blood testing in the current standard of care is often inconvenient and time-consuming. It relies on a centralized laboratory model leveraging laboratories that process blood samples on numerous analyzers spanning multiple assay types^6^. In addition, preanalytical challenges, including incorrect sample storage, handling errors, and prolonged time lag, can introduce the potential complication of skewed diagnostic blood test results, undermining accurate patient diagnoses and treatment decisions. To support testing on the various analyzers, a large blood draw into multiple collection tube types is required. Patients typically need to set up a separate appointment to complete the blood draw, sometimes at a different facility from the treating physician’s office. The samples are then transported to a centralized laboratory for testing, while patients must wait up to a week for their results to be reported back to them. In addition, patients must follow up with a separate appointment with their health care provider to discuss the results in person, especially if the test results are abnormal^7^. This cumbersome process, which is both fragmented and time-consuming^8^, is a key factor for why 40% of patients do not follow up on testing orders^9^. Poor compliance in adherence to prescribed blood test orders and follow-up on blood test results can have major consequences in patient care, including missed diagnoses and suboptimal patient outcomes^7^.

One avenue to improve patient compliance with blood test orders is the adoption of point-of-care (POC) testing systems^10^. POC devices make testing accessible when and where a test is needed, removing many of the pre-analytical challenges involved in centralized testing^6^ and offering shorter turnaround time^6,11^, which in turn enables real-time physician review of the results during the patient’s visit. Regrettably, despite these advantages, the currently available POC analyzers have significant limitations^12^ including restricted test menus, limited support for multiple assay types, and a lack of the accuracy and/or precision afforded by large central laboratory analyzers^12,13^. These limitations prohibit POC devices from replacing the centralized blood testing status quo when on-demand testing is desired.

To address the growing need for accessible, accurate and precise POC testing^14,15,17^ in support of timely clinical decision-making^11,16^ and improve patient outcomes^11,18^, Truvian is developing a fully automated and integrated benchtop device. The Truvian platform utilizes a small (300 μL), heparinized blood sample and is able to simultaneously deliver results for the most commonly ordered routine blood tests, inclusive of comprehensive metabolic panel (CMP) analytes, complete blood count (CBC) with differential, lipid panel, hemoglobin A1c (HbA1c), and thyroid stimulating hormone (TSH) tests in a single test panel. The multi-modal design of the Truvian platform obviates the need for multiple platforms while providing flexibility for future test panel expansion. In short, the Truvian platform presents the unique opportunity to move routine health monitoring to the POC setting from the central laboratory model.

Internal studies together with field testing during development are critical to understand baseline performance across multiple assay types, provide a roadmap for improvements needed, and gain valuable feedback on workflow from untrained operators in a real-world setting. This paper describes the Truvian platform, and its current performance compared against established central laboratory platforms. To assess the performance of the Truvian platform, a multi-site performance study was completed at Truvian’s headquarters in San Diego, California, and at an independent clinical trial site in the Pacific Northwest. The study assessed the concordance of the Truvian platform to central laboratory platforms, while also characterizing the precision, linearity, sensitivity, and tolerance of Truvian assays to interference from hemolytic, icteric, and lipemic sources.

## METHODS

### Truvian Platform and Consumables

The Truvian platform is an automated and integrated compact blood testing device in late-stage development. The platform can simultaneously perform clinical chemistry, immunoassay, and hematology assays with only 300 μL of blood. In addition to the small sample volume required, the Truvian platform can perform all assays with blood from a single blood collection tube type. As opposed to central laboratory platforms, it does not need an experienced technician, upfront pre-analytical processing, significant floor (or bench) space, custom plumbing or electrical power set up. The system has limited infrastructure needs – approximately 1ft by 2ft of countertop space, access to a standard power outlet, and internet connectivity is optional. The Truvian platform (Figure 1a) uses two unique, single-use consumables, a disc (Figure 1b) and a support pack (Figure 1c). These disposables provide all necessary reagents and materials to perform all assays in the panel and act as a waste container which is disposed of at the end of the run. The disc contains a plasma separation feature, a hematocrit feature, and universal microwells where assay reagents are dried and stabilized for room temperature storage. The support pack includes a blood sample holder, pipette tips, additional assay reagents, reagent preparation wells, and a monolayer for hematology analysis. The platform uses a high precision pipettor to handle sample and liquid reagents, an on-board centrifugation apparatus to separate whole blood into plasma, and a closed-loop thermal control system to provide precise temperature regulation required for various assays. Four optical modules collect assay data to provide simultaneous and automated results.

**Figure 1.**
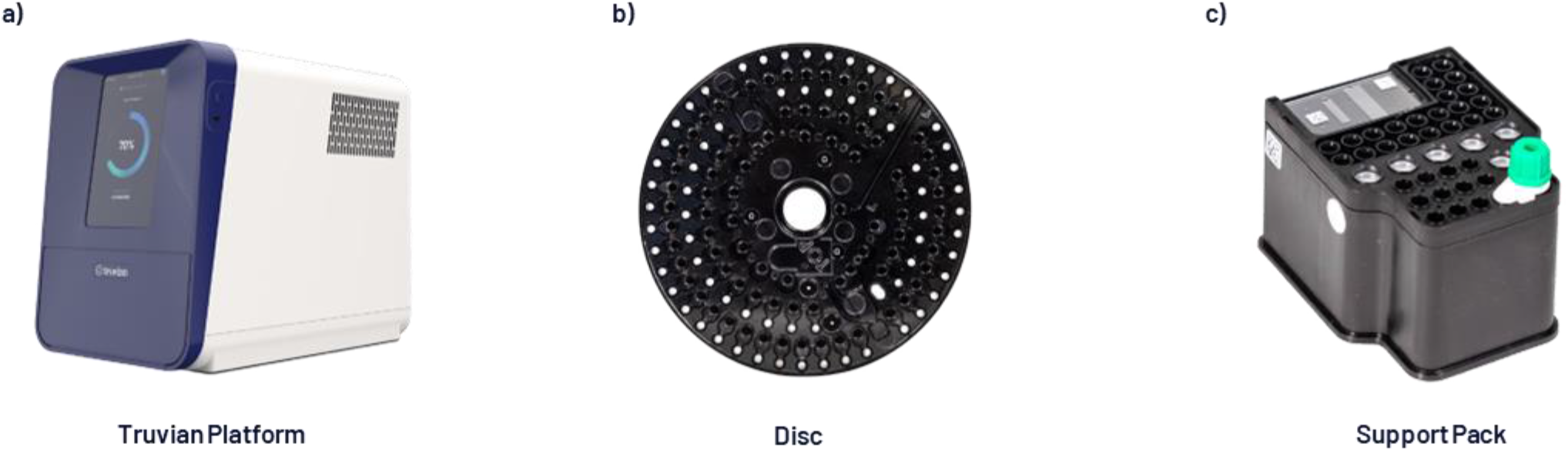
(a) Truvian Platform: an automated multi-modal blood testing analyzer measuring H = 17.25”, L = 20”, W = 12.5”, (b) single-use disc consumable that contains microwells pre-filled with dried chemistries, plasma separation features, and hematocrit channel, (c) single-use support pack that holds the blood sample and contains pipette tips, buffers, reagent preparation wells, and a monolayer.

The four optical modules include:

1. A cell imager to collect high resolution brightfield and fluorescent images for hematology tests.
2. A spectrophotometer to measure the light absorbance through the microwells for clinical chemistry tests.
3. A confocal fluorescent laser scanning module for bead-based immunoassays.
4. A CCD camera for collecting assay readings in addition to quality control images.

The Truvian platform is designed for ease of use and is factory calibrated. To perform a test, an operator inserts a blood sample into the support pack, places it into the platform with the disc, and follows a guided user interface (UI) workflow (Figure 2). From there, the device processes the blood and performs the panel of tests, which are measured with the optical modules described above. Proprietary onboard algorithms compute the results in real-time and a report is shown on the device through the touchscreen interface at the end of each run. Upon completion of the run, the report can be saved and/or printed and the consumables are then automatically ejected for disposal. If configured with internet access, the report can be transmitted to the cloud for integration into a laboratory information system (LIS) or an electronic medical record (EMR).

**Figure 2.**
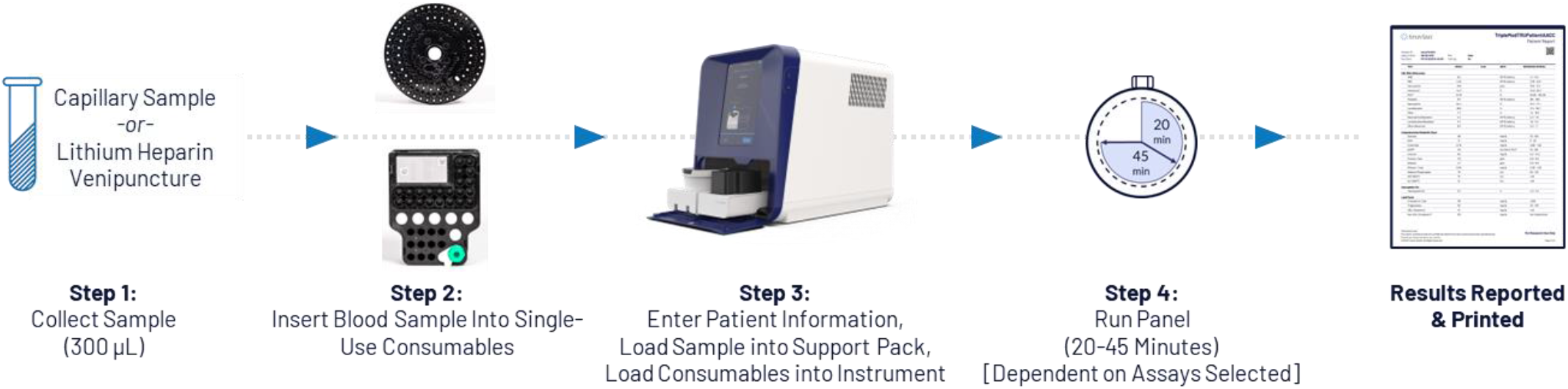
The Truvian platform is fully integrated and automated with no upfront processing or pipetting steps required. Step 1: Collect minimum 300 μL whole blood sample, Step 2: Insert the sample (300 μL tube of Li-Hep blood) into the Support Pack, Step 3: Enter patient information and place single-use Disc and Support Pack into the device, Step 4: Start the panel. At the end of the run, panel results are displayed on the device and can be printed. The consumables are then ejected to be disposed.

### Study Design

The multi-site performance evaluation study was conducted 1) to characterize a multi-modality panel on the Truvian platform, 2) to test the platforms at an external clinical trial site to simulate real-world settings, and 3) to determine the concordance of results obtained from Truvian’s platforms compared to gold-standard central laboratory platforms.

The precision and panel characterization studies including linearity, sensitivity (LoB/LoD/LoQ) and interfering substances testing were performed at Truvian headquarters in San Diego, California. The method comparison study was conducted at two locations: Truvian headquarters in San Diego, California, and an independent clinical trial site in the Pacific Northwest. Altogether, the studies were performed across 10 Truvian platforms using a multi-modality panel that included a total of 32 tests: albumin (ALB), alkaline phosphatase (ALP), alanine aminotransferase (ALT), blood urea nitrogen (BUN), calcium (CA), creatinine (CRE), estimated glomerular filtration rate (eGFR), glucose (GLU), glycated hemoglobin A1c (HbA1c), high density lipoprotein (HDL), low density lipoprotein (LDL), non-HDL cholesterol (non-HDL), total cholesterol (CHOL), total cholesterol/HDL ratio (CHOL/HDL), total bilirubin (TBIL), total protein (TP), triglycerides (TRIG), thyroid stimulating hormone (TSH), very low density cholesterol (VLDL), red blood cell count (RBC), white blood cell count (WBC), platelet count (PLT), hematocrit (HCT), hemoglobin (HGB), mean corpuscular volume (MCV), lymphocyte count (LYMPH), lymphocyte percentage (LYMPH%), neutrophil count (NEUT), neutrophil percentage (NEUT%), other WBC count (OTHER), other WBC percentage (OTHER%). Specifically, LDL was calculated using the Friedewald equation and eGFR was calculated using the CKD-EPI creatinine equation (2021).

At Truvian’s headquarters, the Truvian platforms were operated by laboratory staff who are familiar with the operations of the platform. At the independent clinical trial site in the Pacific Northwest, the Truvian platforms were operated by untrained operators to test the usability of the platform in a real-world setting. The operators were given a brief written protocol and a demonstration of the operation prior to the study.

These studies were conducted in June and July of 2023. The method comparison study protocols were submitted, reviewed, and approved by an independent Institutional Review Board.

### Precision Study

The precision study, based on CLSI EP05-A3 guidelines, was used to evaluate repeatability and reproducibility using commercially-available controls for clinical chemistry, hematology, and immunoassay. Fresh vials of control material (Bio-Rad and R&D Systems) were used during five days of testing. Four replicates per level were run each day on each of the three platforms, producing a total of 180 measurements per analyte for a total of 4500 test results. Reproducibility was evaluated for each level and analyte. The reproducibility CV was calculated as the sum of between-platform, between-day, and within-run (repeatability) variances.

### Linearity

Linearity was evaluated using a study design based on the CLSI EP06 guidelines. These studies were performed utilizing contrived samples (9-15 levels varying per assay) across clinically relevant ranges, spanning the Analytical Measuring Range (AMR) for each test. For clinical chemistry, a total of 142 runs over 5 days were performed on 5 instruments. Four different linearity sample sets were generated: the HbA1c linearity panel (10 levels + 6 intermediate blends), the Hemoglobin linearity panel (10 levels), the Total Bilirubin linearity panel (9 levels), and the plasma linearity panel, for additional assays: Alb, ALP, ALT, AST, BUN, Ca, Chol, Creat, Gluc, HDL, TP, Trig) (15 levels). For hematology, RBC/HCT, WBC, and PLT were run with 3 separate panels across 1 day each and with up to 8 instruments per day. Between 9 to 11 linearity levels spanning the AMR were included for each analyte.

### Sensitivity (LoB/LoD/LoQ)

Limit of Blank (LoB), limit of detection (LoD), and limit of quantitation (LoQ) were determined using a study design based on the CLSI EP17 guidelines to determine the detection capabilities for each assay. The results from these studies helped establish the functional range of the Truvian platform across all three detection methods. For clinical chemistry assays, the LoB samples were water and saline, the LoD and LoQ material were donor samples contrived to meet target levels. For clinical chemistry LoB/LoD, 80 runs were performed across 4 instruments for plasma-based assays, and 40 runs for blood-based assays across 4 days of testing. LoB/LoD were tested with 4 blanks, and 4 low-level samples. For LoQ, 24 runs were performed for plasma-based assays, and 12 runs for blood-based assays for each of the 4 LoQ sample levels over 3 days, across 4 instruments. Regarding hematology assays, for WBC and PLT, 20 replicates were run for each assay in 1 day of testing for 1 ultra-low sample (LoB). Additionally, for each test, two samples per day were run for a total of 10 runs per sample for each of the 4 low-level samples (LoD), and 1 level sample was created for LoQ that was run for 20 replicates. A total of 8 instruments were used for WBC LoB/LoD/LoQ, and 9 instruments were used for PLT LoB/LoD/LoQ. For the TSH assay, 59 runs were performed across 4 days for LoB, and 57 runs across 3 days for LoD/LoQ on 2 instruments. A total of 5 LoD/LoQ samples were prepared by adding recombinant human TSH into affinity stripped TSH free matrix. TSH LoB samples were prepared from unique TSH affinity stripped or Charcoal Stripped, Fumed Silica Delipidized Serum.

### Interference

Sample quality assessment (SQA) was performed using a study design based on CLSI EP07 guidelines, to determine levels of common interferants (hemoglobin, bilirubin (conjugated and unconjugated), and lipids), often referred to as HIL, in samples. Tolerance to HIL endogenous interferences was established for all assays and compared to FDA-cleared predicate methods. For clinical chemistry assays, dose-response interference testing was performed with interferant-spiked contrived plasma up to 1000 mg/dL lipids (Intralipid), 800 mg/dL hemoglobin (donor-derived hemolysate), and 45mg/dL bilirubin (conjugated and unconjugated). Interferant concentrations were estimated using serum index measurements on the Roche Cobas. Tests were run across multiple instruments and analyte recovery was determined in comparison to control plasma spiked with the corresponding interferant diluent. Statistically significant interference was determined using a 2-sided t-test. For hematology, screening testing was performed with 4 interferants: (Bilirubin C, Bilirubin F, Intralipid, and Hemoglobin) at the maximum concentrations and unspiked control with 5 replicates. Additionally, dose-response testing was performed for 5 different levels with 5 replicates to identify threshold of each interferant for hematology analytes. For TSH, a total of 19 runs were performed to compare results between two control plasma matrix samples with three contrived high level interferant samples (targeting 1000 mg/dl Intralipid, 800 mg/dl hemoglobin, and 45 to 57 mg/dL bilirubin (conjugated and unconjugated). Analytical thresholds were within ±10% of the control material value to be acceptable as having ‘no significant effect’ on TSH levels.

### Method Comparison Study

The multi-site method comparison study was performed to compare the Truvian platform’s results to those obtained from Roche Cobas (for chemistry and immunoassay) as well as the Sysmex XN and pocH-100i (for hematology) in normal donors and patients with chronic disease. A total of 237 donors’ results were evaluated.

Donors were consented under IRB approved protocol PL-007-2023-006 or 0012022 (IRB Registration: IRB00000533), with each participant donating three tubes of venous blood. One tube of EDTA anticoagulated blood was collected for analysis on the Sysmex hematology analyzer. One lithium heparin (Li-Hep) anticoagulated blood in a gel-separator tube was collected and centrifuged to isolate plasma for the Roche Cobas. The third tube contained LiHep anticoagulated blood, which was run on the Truvian platform within one hour of collection.

The EDTA and Li-Hep samples were either analyzed in-house at Truvian (approx. 40%) on comparator devices or sent to a central laboratory (approx. 60%) for testing. Samples sent to the central laboratory were processed following central laboratory instructions and stored at 2-8 °C until courier collection. An estimate of 49% of the samples were from apparently healthy donors, 43% from patients with chronic diseases such as liver disease, heart disease, kidney disease, diabetes, and thyroid disorder, as well as 8% contrived samples to assess Truvian’s performance across an extended analytical measuring range and around medical decision points.

Contrived samples were prepared for selected tests found to have inadequate coverage of the measuring range. High and low samples were contrived for ALB, TP, and CA by spiking fresh donor whole blood with concentrated analyte stocks or diluting the whole blood sample with saline, respectively. These samples were included for other assays when found to be commutable with donor samples.

### Data Analysis

Analyse-It software was used to assess the precision study data per CLSI EP05-A3 guidelines. Measurement system analysis was performed using a 2-factor model in which the testing day was nested within each platform. A 2-sided confidence interval of the standard deviation was determined for each analyte at the 95% level. The reproducibility was calculated as the sum of between-platform, between-day and within-run (repeatability) variances and reported as standard deviation and %CV.

The analysis of linearity datasets was performed using Analyse-It software, as per CLSI EP06 guidelines. Allowable deviation from linearity criteria for most assays was set at the standard recommended ±10%. However, for some assays different criteria were used based on comparator device performance claims.

Regarding sensitivity data analysis, Analyse-It software was also utilized, following CLSI guidelines. For the analysis of LoQ data, a CV % threshold was set as acceptance criteria, varying for the different assays, with reference to FDA-cleared predicates.

For the method comparison study, bias was estimated using Passing-Bablok or Deming regression and Bland-Altman difference/relative difference measures. Estimates of correlation and limits of agreement were also calculated. Confidence intervals for regression coefficients were determined using the Bootstrap or Jackknife methods. Bland-Altman difference/relative difference estimates were compared to CLIA, NCEP, or NGSP allowable total error criteria.

For interference testing, analyte recovery was determined in comparison to control sample spiked with the corresponding interferant diluent; statistically significant interference was determined using a 2-sided t-test according to CLSI EP07.

## RESULTS

Truvian instruments demonstrated > 95% run reliability at both the Truvian HQ and the Pacific Northwest clinical trial site. Run reliability is defined as the percentage of runs that completed and generated results successfully. Assay performance measured at the clinical trial site was also consistent with data collected at Truvian.

### Precision Study

Results from the precision study met specifications for all measured tests evaluated. Results are summarized in Table I. Of the 4500 total datapoints, there was one single outlier excluded from the results. This outlier was from the RBC mid-level control runs, for which a root cause was identified. Since executing this study, a QC metric has been developed and implemented to suppress reporting of this type of outlier in the future.

**Table 1.**
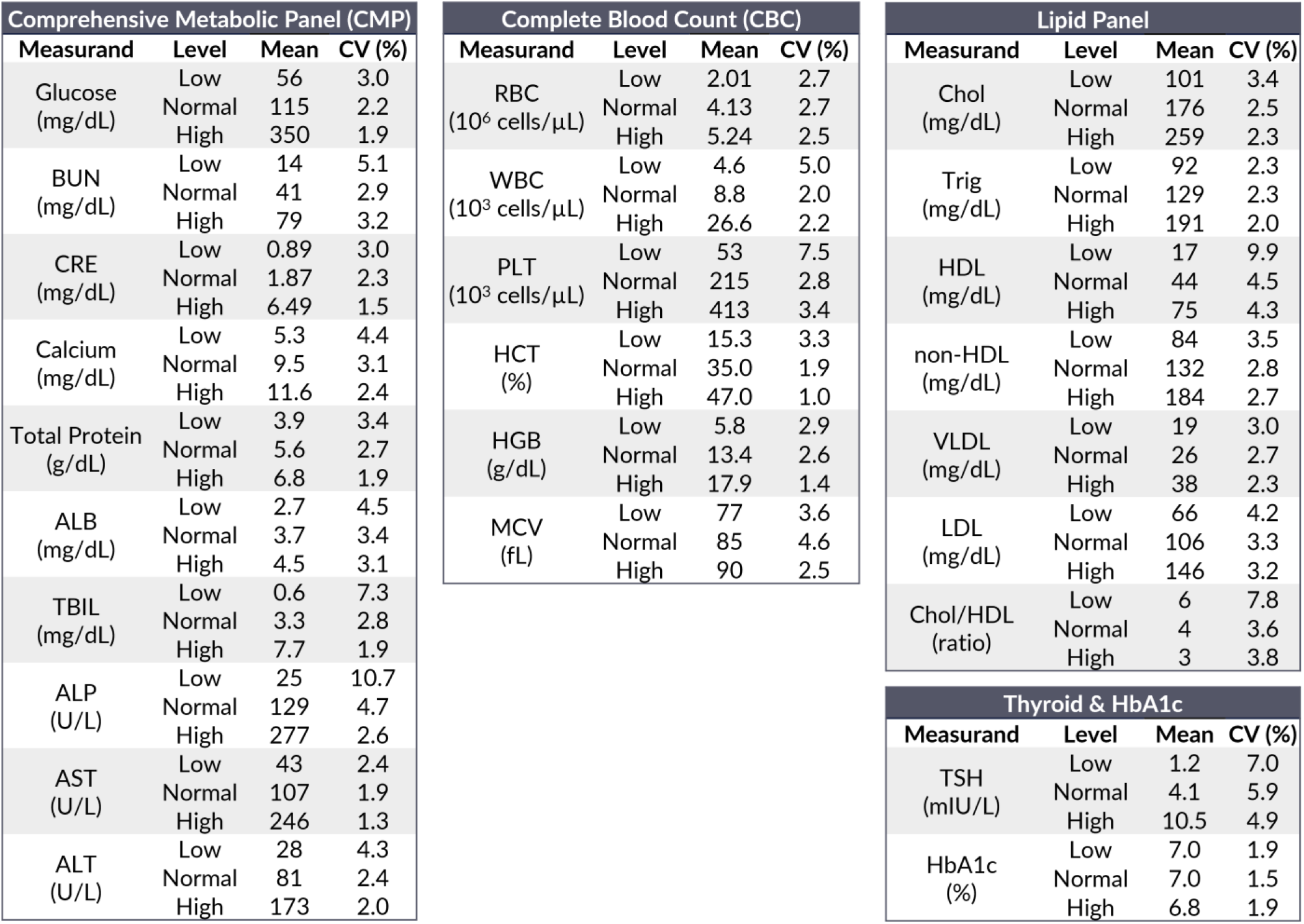
Precision Study Results.

All tests showed consistent reproducibility, demonstrating stable performance across multiple instruments during the testing period.

### Linearity, Sensitivity and Specificity

To evaluate linearity and sensitivity (Figure 3), a subset of tests was selected to highlight performance across all detection methods including immunoassay, endpoint and kinetic clinical chemistry, immunoturbidimetry, and cell counting tests. All tests evaluated demonstrated sensitivity comparable to other commercially available point of care devices and maintained linearity across the entire dynamic range. Importantly, TSH demonstrated a linear range of 0.06 to 45.7 mIU/L with an LoB of 0.01 mIU/L, an LoD of 0.04 mIU/L and an LoQ of 0.06 mIU/L, which is consistent with 3^rd^ generation TSH tests.

**Figure 3.**
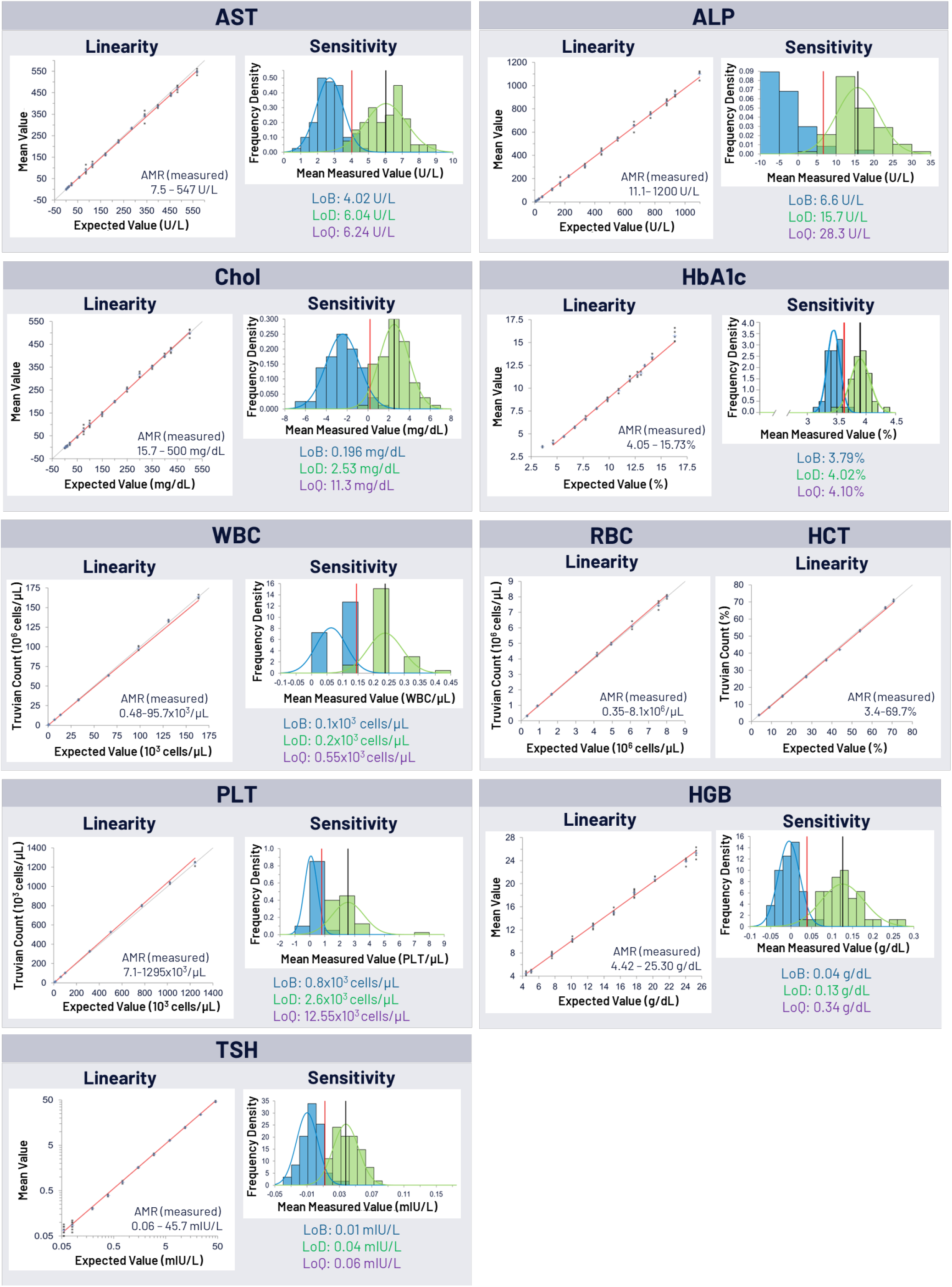
The Truvian Platform shows good linearity and sensitivity across analyte measuring range (AMR).

The tolerance of various tests to common endogenous interfering conditions (hemolysis, icterus and lipemia) was also evaluated (Table 2a, 2b, and 2c). The levels of tolerance to hemolysis, icterus, and lipemia for tests on the Truvian platform are comparable to those on predicate devices. The tests evaluated tolerated a range of 50 mg/dL to 1000 mg/dL of hemoglobin with TBIL, ALP, and AST being the most sensitive while WBCs and platelets were the most tolerant. The majority of assays evaluated for sensitivity to icterus showed tolerance between 30 – 45 mg/dL bilirubin. The absorbance-based tests glucose, creatinine, total protein, cholesterol, and triglycerides were the most sensitive and could only tolerate between 5 -15 mg/dL bilirubin. Lipemia evaluation showed good tolerance (500 to 1000 mg/dL Intralipid) across all analytes evaluated.

**Table 2A.** Thresholds for Hemolysis Interference.

| Hemolysis Index : Hemoglobin (mg/dL) |  |  |  |  |  |
| --- | --- | --- | --- | --- | --- |
| CMP |  | CBC |  | Lipid Panel |  |
| Glucose | >500 | RBC | 250 | Chol | 175 |
| BUN | >800 | WBC | 1000 | Trig | 100 |
| CRE | >500 | PLT | 1000 | HDL | 400 |
| Calcium | >500 | HCT | 900 | Thyroid & HbA1c |  |
| Total Protein | 800 | HGB | N/A | TSH | >800 |
| ALB | >800 | Lymphocytes | 1000 | HbA1c | N/A |
| TBIL | 50 | Neutrophils | 1000 |  |  |
| ALP | 50 | Other | 1000 |  |  |
| AST | 50 |  |  |  |  |
| ALT | 400 |  |  |  |  |

**Table 2B.** Thresholds for Icterus Interference.

| Icterus Index : Bilirubin Conjugated / Unconjugated (mg/dL) |  |  |  |  |  |
| --- | --- | --- | --- | --- | --- |
| CMP |  | CBC |  | Lipid Panel |  |
| Glucose | 10 / >20 | RBC | 40 / 36 | Chol | 5 / 10 |
| BUN | >45 / >45 | WBC | 40 / 40 | Trig | 5 / 7.5 |
| CRE | >10 / >10 | PLT | 40 / 18 | HDL | >20 / 20 |
| Calcium | >30 / >30 | HCT | 40 / 20 | Thyroid & HbA1c |  |
| Total Protein | 15 / 10 | HGB | >45 / >45 | TSH | >45 / >45 |
| ALB | >45 / >45 | Lymphocytes | 40 / 40 | HbA1c | >45 / >45 |
| TBIL | N/A | Neutrophils | 40 / 40 |  |  |
| ALP | >45 / >45 | Other | 40 / 40 |  |  |
| AST | 45 / 20 |  |  |  |  |
| ALT | 45 / 20 |  |  |  |  |

**Table 2B.** Thresholds for Lipemia Interference.

| Lipemia Index : Intralipid (mg/dL) |  |  |  |  |  |
| --- | --- | --- | --- | --- | --- |
| CMP |  | CBC |  | Lipid Panel |  |
| Glucose | 500 | RBC | 720 | Chol | >1000 |
| BUN | >1000 | WBC | 800 | Trig | N/A |
| CRE | >1000 | PLT | 800 | HDL | 1000 |
| Calcium | 750 | HCT | 800 | Thyroid & HbA1c |  |
| Total Protein | >1000 | HGB | >1000 | TSH | >1000 |
| ALB | >1000 | Lymphocytes | 800 | HbA1c | >1000 |
| TBIL | 750 | Neutrophils | 800 |  |  |
| ALP | >1000 | Other | 800 |  |  |
| AST | 300 |  |  |  |  |
| ALT | 300 |  |  |  |  |

### Method Comparison Study

In the method comparison study, results from a total of 237 samples consisting of 49% apparently healthy, 43% donors with chronic diseases, and 8% contrived samples were evaluated for concordance and equivalency using regression (Passing-Bablok or Deming) (Figure 4a, b & c) and Bland-Altman analyses (Table 3), comparing results from the Truvian platform to those from either the Roche Cobas (chemistry and immunoassays) or Sysmex XN or pocH-100i (hematology).

**Figure 4A.**
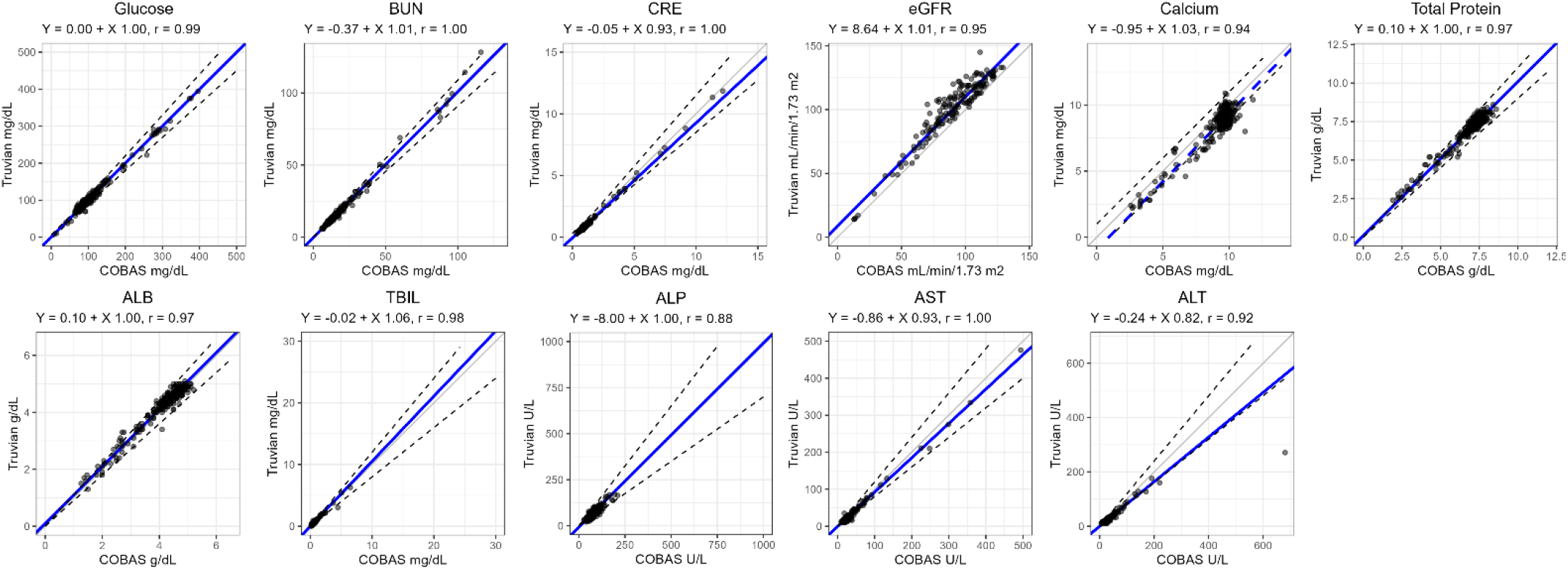
Method Comparison Truvian platform and Roche for partial Comprehensive Metabolic Panel (CMP)

**Figure 4B.**
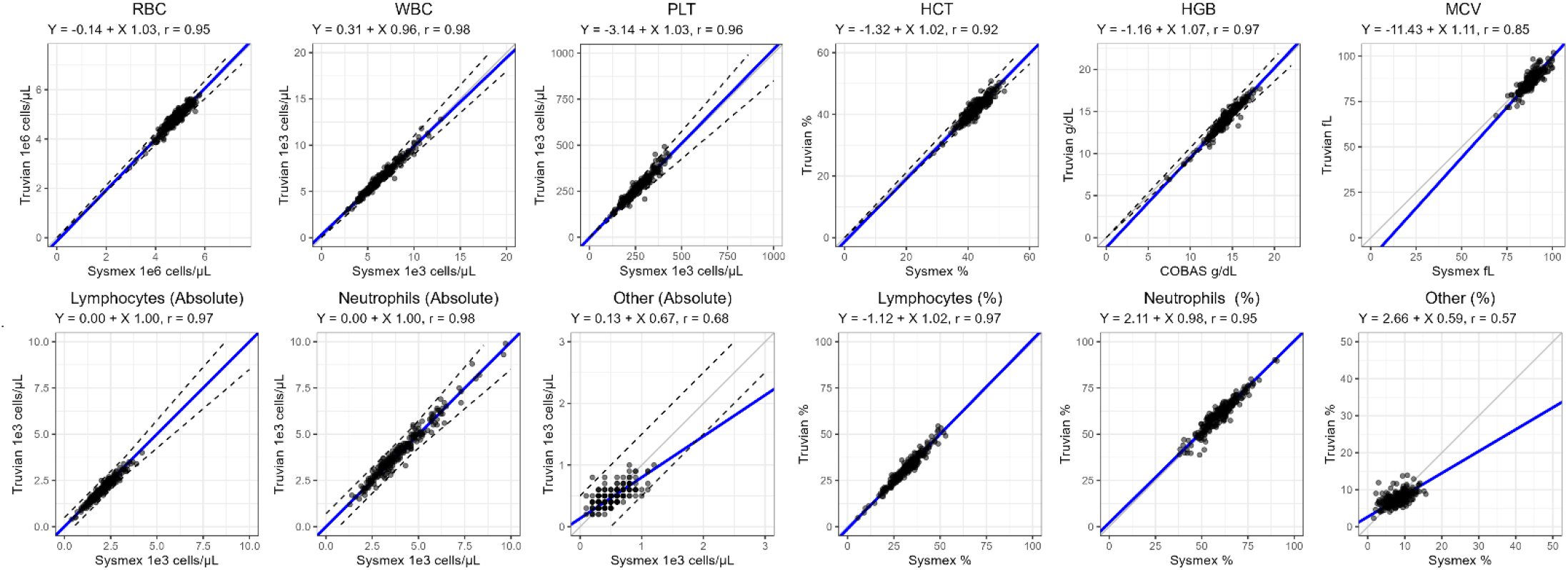
Method Comparison Truvian Platform and Sysmex for Complete Blood Count (CBC)

**Figure 4C.**
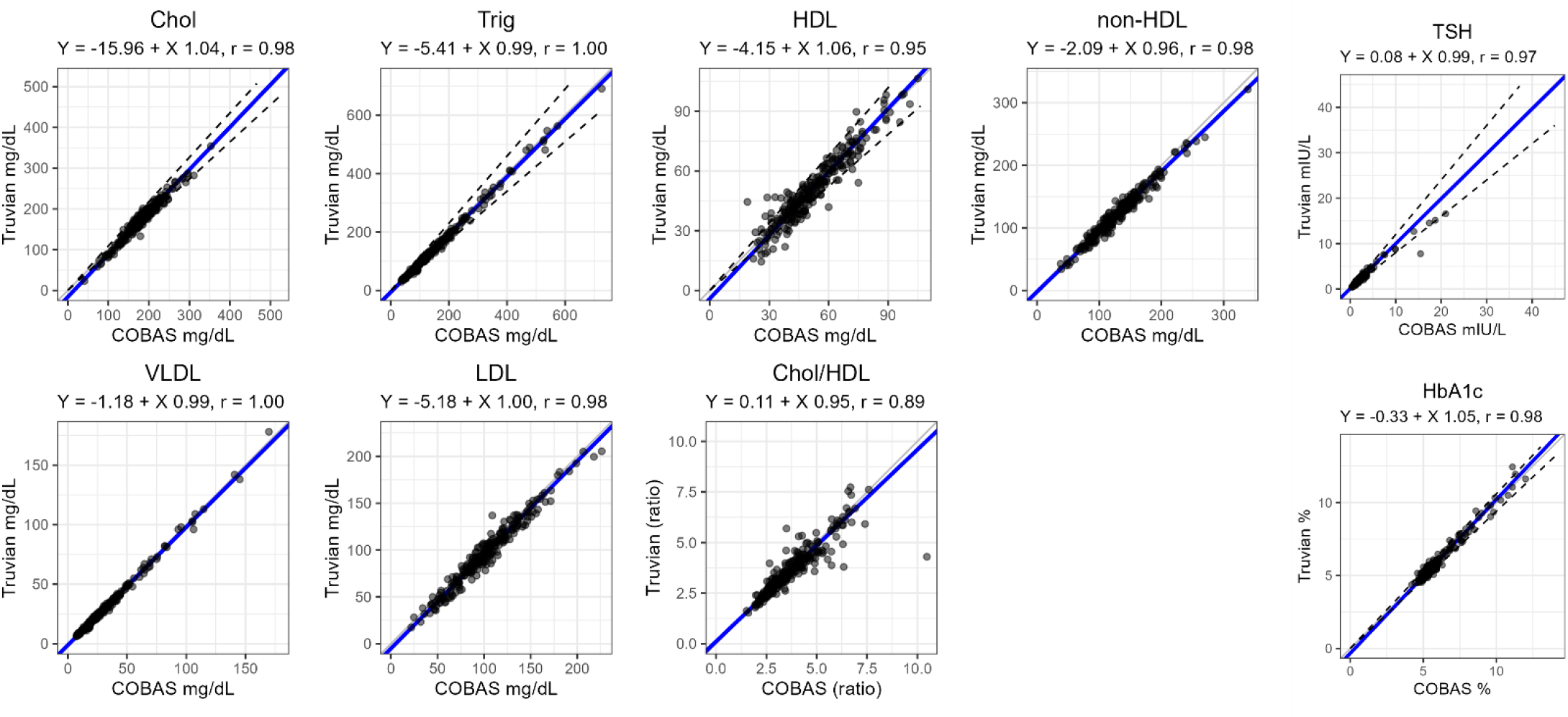
Method Comparison the Truvian Platform and Roche for Lipid Panel, Thyroid Panel and HbA1c

The regression analysis presented in Figures 4a, 4b, 4c, and Table 3, showed strong concordance between the Truvian platform and central laboratory results for the majority of tests with no observed differences between internally and externally collected data. ALB, TP, and CA tests had insufficient coverage of the AMR. This was addressed by incorporating contrived samples to expand coverage of the AMR.

## DISCUSSION

This multi-site study evaluated the precision, detection capability, and accuracy the Truvian platform, which is in late-stage development. The main goals for conducting this study were to understand the performance of the Truvian platform in a real-world setting and to evaluate precision, sensitivity, specificity, and accuracy of the Truvian platform against performance specifications. Performance specifications were established based on the performance of predicate devices as well as CLIA guidelines.

The high run reliability (> 95%) during the course of the study demonstrated a consistent and dependable performance of the core hardware design. The data presented here demonstrated that the bench top, multi-modal Truvian platform is capable of producing accurate, precise, sensitive, and specific results at the point-of-action^™^, and comparable to large central laboratory analyzers.

The precision study showed that the Truvian platform produced repeatable and reproducible results, demonstrating the interoperability of multiple Truvian platforms. Notably, there was one single outlier identified across the 4500 datapoints through statistical analysis and excluded from the RBC mid-level control runs. Since testing, we have addressed this type of outlier through the development of a QC (Quality Control) metric included in our algorithm to detect anomalous results.

The linearity and sensitivity studies showed that the Truvian platform is consistent across the dynamic range of all tests evaluated with good tolerance of common interfering conditions (hemolysis, icterus, lipemia), comparable to predicate devices. Notably, results from these studies demonstrate that the Truvian TSH assay is on par with 3^rd^ generation TSH assays with an LoQ of 0.06 mIU/L and dynamic range of 0.06 mIU/L to 45.7 mIU/L.

The method comparison study showed encouraging results with good concordance between Truvian platform and the central laboratory results as assessed by Passing-Bablok or Deming regression and Bland-Altman analyses in 237 donor samples along with banked abnormal patient samples and contrived samples.

This study provides strong evidence of the core viability of the Truvian platform, and the results will be further strengthened as part of ongoing development prior to formal verification and validation and submission to the FDA.

Since this study concluded, we have incorporated panel workflow enhancements to generate the full suite of results under 30 minutes without compromising performance. This change will make it possible to release the full panel of results that takes 1-3 days at a central laboratory to under 30 minutes on the Truvian platform. This optimized time-to-result (TTR) panel will incorporate new reagent formulations that improve performance for a number of tests (CA, CRE, ALP and ALT) in conjunction with on-board sample quality checks to automatically detect and flag results for the presence of endogenous interference. Ongoing and future studies with pathological and contrived samples will leverage these platform improvements. Additionally, calibration and value assignment strategies will be further refined and validated using international reference standards or methods where possible.

The Truvian platform is a novel, fully automated, benchtop blood testing platform which aims to disrupt how blood testing is performed and experienced in today’s healthcare system. Our preliminary data strongly indicate that our multi-modal platform can deliver accurate results comparable to three different FDA-cleared central laboratory devices, all from a single sample tube. This innovative, first-in-class technology brings central lab-level performance right to the point-of-action^™^ and stands to facilitate more meaningful interactions between patients and providers. Continuing efforts are underway to complete development of the platform in order to initiate formal verification and validation studies which will result in a formal submission to the FDA.

## Data Availability

All data produced in the present study are available upon reasonable request to the authors.

